# Genetics identifies obesity as a shared risk factor for co-occurring multiple long-term conditions

**DOI:** 10.1101/2024.07.10.24309772

**Authors:** Ninon Mounier, Bethany Voller, Jane AH Masoli, João Delgado, Frank Dudbridge, Luke C Pilling, Timothy M Frayling, Jack Bowden, the GEMINI Consortium

**Author notes:** Corresponding author: Prof Jack Bowden.

## Abstract

**Background:** Multimorbidity, the co-occurrence of multiple long-term conditions (LTCs), is an increasingly important clinical problem, but often little is known about the underlying causes. Observational studies are highly susceptible to confounding and bias, and patients with multiple LTCs are usually excluded from randomised controlled trials. We investigate the role of a potentially critical multimorbidity risk factor, obesity, as measured by body mass index (BMI), in explaining shared genetics amongst 71 common LTCs.

**Methods and Findings:** In a population of northern Europeans, we estimated unadjusted pairwisegenetic correlation, 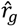, between LTCs and partial genetic correlations after adjustment for the genetics of BMI, 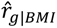. We compared these correlations using a bespoke block-jackknife approach to assess whether differences between the estimates were statistically meaningful. We then used multiple causal inference methods to confirm that BMI causally affects not only individual LTCs, but also their co-occurrence. Finally, we attempted to quantify the population-level impact of intervening and lowering BMI on the prevalence of 15 key common multimorbid LTC pairs. Our results showed evidence that BMI partially explains some of the shared genetics for 740 LTC-pairs (30% of all pairs considered). For a further 161 LTC-pairs, the genetic similarity between the LTCs was entirely accounted for by BMI genetics. This list included diabetes and osteoarthritis: 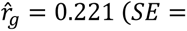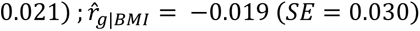, as well as those involving LTCs from the same broad family, or ‘domain’, such as gout and osteoarthritis: 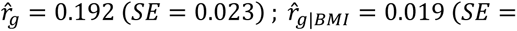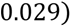. Causal inference methods confirmed that higher BMI acts as a common risk factor for a subset of these pairs, and therefore BMI-lowering interventions would reduce the prevalence of these pairs of LTCs. For example, we estimated that a 1 standard deviation or 4.5 unit decrease in BMI would result in 17 fewer people with both chronic kidney disease and osteoarthritis per 1000 who currently have both LTCs.

**Conclusions:** Our genetics-centred approach shows that obesity is an important mechanistic cause of many shared long-term conditions. We identify LTC pairs for which obesity is the predominating shared risk factor, and cases where it is one of the several shared risk factors involved. Our method for calculating full and partial genetic correlations is published as an R package *{partialLDSC}* for use by the research community.

## Introduction

Multimorbidity, defined as the coexistence of two or more long-term conditions (LTCs), is an important public health challenge. The prevalence of multimorbidity differs across geographic regions, age groups and between genders. It is also higher in more deprived individuals and is often associated with lower quality of life and increased healthcare costs [1]. Many observational studies have focused on defining and measuring multimorbidity [2,3]. However, disparate definitions have led to variations in its characterisation [4]. For example, a thorough definition of chronicity and a list of chronic LTCs have been proposed by [5] and new approaches to cluster LTCs and identify patterns of multimorbidity have been developed [6,7]. These approaches have limitations, as they often involve single datasets and modalities, and cluster identification might differ depending on the algorithm [8].

Investigating the role of common epidemiological risk factors for multimorbidity is vitally important, as it can provide a better understanding of the mechanisms underlying the co-occurrence of LTCs and help to develop efficient prevention strategies. For example, using observational data to study the relationship between socio-economic status and multimorbidity, it has been shown that lower education level and higher deprivation were associated with increasing risk of multimorbidity [9]. A large multi-cohort prospective study provided evidence of associations between obesity and a wide range of LTCs, as well as with the number of LTCs developed, highlighting the potentially important role of obesity in multimorbidity [10].

We have previously assessed the genetic similarity between 2546 pairs of LTCs [11], allowing us to re-examine associations from observational epidemiology. Using genetic predictors of a trait, rather than observational measures, reduces the impact of inherent issues such as confounding, measurement error and reverse causation [12]. These issues are particularly problematic for studies of multimorbidity. Over the last decade, genetic approaches have also greatly benefited from the increase in sample sizes, and methods that enable data from more than one cohort to be incorporated into the analysis, enabling researchers to work with data linking tens of thousands of cases for common LTCs from a large range of health exposures. This genetics-centred approach has been used to estimate the costs of obesity on healthcare systems [13]. Our previous work showed evidence of widespread genetic correlation across LTCs, with obesity being highly genetically correlated with a broad range of LTCs [11]. Depending on context, obesity can be considered a risk factor for multiple LTCs or an LTC in its own right. In this work, we focus on using statistical genetics methods to quantify the role of obesity as a common risk factor for multimorbidity.

Genetic correlation is driven by pleiotropy, when a genetic locus affects several traits, and can reflect different scenarios: a direct relationship between the two traits (vertical pleiotropy – when the two traits are part of a causal cascade), a common biological process or the effect of a common risk factor on both traits (horizontal pleiotropy – when the two traits have no direct effect on each other) [14] (Supplementary Figure 1). Often, genetic correlations are driven by a combination of both vertical and horizontal pleiotropy [15]. We propose an approach to help differentiate these mechanisms in multimorbidity, focussing on obesity as a well-known risk factor for several long-term conditions [16]. We chose the most common, general clinical measure of obesity for this investigation: body mass index (BMI).

We use data from the GEMINI collaborative for 71 long-term common LTCs comprising 2485 distinct pairs. We propose an approach to compare pairwise unadjusted genetic correlations to partial genetic correlations that accounts for BMI genetics, formally testing whether BMI explains a portion of the genetic correlation for a given LTC pair. We also applied causal inference methods to elucidate the causal (biological) mechanisms through which BMI affects the genetic correlation between LTCs, quantifying its pivotal role as a common risk factor in multimorbidity.

## Methods

### Data resources

We used GWAS summary statistics, derived from individuals of European descent, for 71 common and heritable LTCs, encompassing 13 distinct disease domains, grouped according to the International Classification of Disease (ICD): such as cardiovascular or respiratory domains [17]. These GWAS data are described in detail in [11]; relevant diagnostic and analytical code is available on the project GitHub pages [https://github.com/GEMINI-multimorbidity]. LTCs were defined by adapting existing diagnostic code lists with input from clinical experts. LTCs were selected for genetic analyses if reaching a prevalence greater than 0.5% in people over 65 in each of two large population-based cohorts in the UK and Spain [18,19]. *Heritability* measures the proportion of phenotypic variance explained by genetics. We estimated this in UK Biobank [20] to identify a subset of LTCs with a genetic basis. Finally, for each condition, we used the largest sample size available by combining GWAS data from up to three sources: UK Biobank, FinnGen, and condition specific consortium data (details available in Supplementary Table 1). For many of these LTCs, the meta-analysed data used represents the largest sample size used to date. We refer to these meta-analysed GWAS summary statistics as "GEMINI summary statistics" for the rest of the paper.

We also used two different sets of GWAS summary statistics for BMI. First, we used the largest data to date (*N* ≈ 700,000), combining results from the GIANT Consortium and UK Biobank [21], to estimate partial genetic correlations. We also used earlier results from the GIANT Consortium (*N* ≈ 340,000 – minimal overlap with the GEMINI summary statistics) [22], to perform causal inference analyses, after ensuring that the partial genetic correlations results were consistent with the ones obtained using the larger dataset.

The GWAS summary statistics for the 71 LTCs, as well as BMI, were pre-processed using the munging function from the LD-score regression (LDSC) [23] and for all LDSC analyses we used LD-scores estimated from the 1000G EUR reference panel.

### Covariance and correlation

*Covariance* measures the extent to which the observed value of one quantity predicts the value of another quantity. For example, in the case of two such quantities – osteoarthritis (OA) and type 2 diabetes (T2D) diagnoses in a study population - a positive covariance would indicate that having T2D increases the probability of having OA. *Correlation* is simply a scaled version of covariance that lies between -1 and 1. If all individuals in a population with T2D also have OA, the two would have a correlation of 1. If all individuals in a population with T2D do not have OA, the two would have a correlation of -1.

### Partial genetic covariance and correlation

In this paper we extensively study the genetic covariance and correlation between two LTCs. That is, the degree to which genetic variants that predict one LTC also predict the other LTC. Furthermore, we focus on understanding how the genetic covariance between two LTCs changes when we remove the genetic effects of a common risk factor.

Using the Schur complement, the partial (or ‘adjusted’) genetic covariance between LTCs *k* and *l* 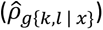, which corresponds to their genetic covariance while holding the genetic effects of the trait *x* (in our case, BMI) constant, can be defined as follows:

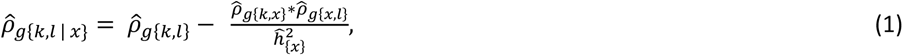

where 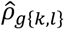 is the genetic covariance between condition *l* and *k*,

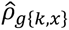 is the genetic covariance between condition *k* and trait *x*,

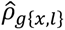 is the genetic covariance between trait *x* and condition *l*,

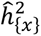 is the heritability estimate for trait *x*.

Genetic covariance and heritability estimates were obtained from GWAS summary statistics using cross-trait LD Score regression (LDSC), as previously described by [24].

Similarly, the partial heritability for LTCs *k* and *l* (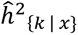 and 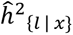, respectively) can be estimated:

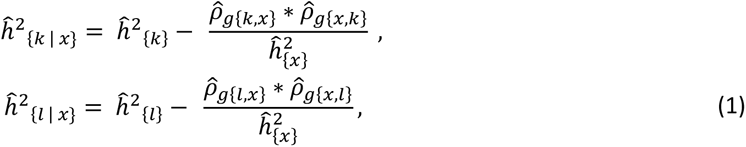

where 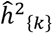 is the heritability estimate for condition *k*,

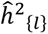 is the heritability estimate for condition *l*.

Finally, the (unadjusted) genetic correlation 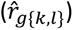 and the partial genetic correlation 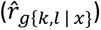 between LTCs *k* and *l* can be derived:

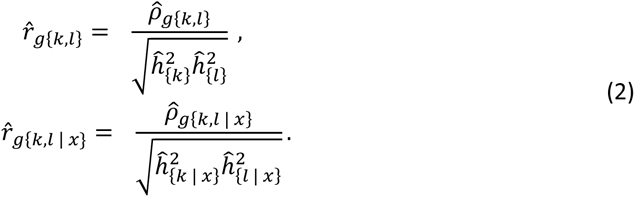

To test if the partial (or adjusted) genetic correlation estimate is different from the unadjusted genetic correlation estimate, the following test statistic was used:

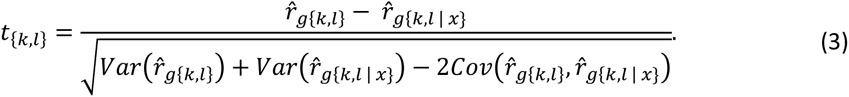

The variance of the unadjusted genetic correlation 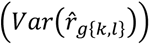, the variance of the partial genetic correlation 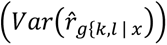 and the covariance between the two 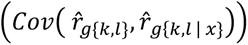 were obtained using a novel block-jackknife approach (see details in Supplementary Note 1). This enabled us to rigorously test for a difference between the two, for the first time.

We estimated partial genetic correlations between all pairs of LTCs, using two different sets of GWAS summary statistics for BMI [21,22]. False discovery rate (FDR) correction (*Q* − *value* < 0.05) was applied for both unadjusted and partial genetic correlations, and the differences between the two, to account for multiple testing. LTC pairs were then first classified according to the difference between the two (FDR-corrected statistically significant difference or not). Significant LTC pairs were then further classified into four categories:

- Both unadjusted and partial genetic correlations were statistically non-significant (FDR of 5%);
- Both unadjusted and partial genetic correlations were statistically significant;
- Only unadjusted genetic correlations were statistically significant;
- Only partial genetic correlations were statistically significant.

Here, we focus on the results with evidence of genetic similarity before and/or after adjustment for BMI. LTC pairs were defined as *within-domain*, if both LTCs belong to the same disease domain, or *cross-domain*, if the two LTCs belong to different disease domains.

Our analysis pipeline has been implemented in a R package - partialLDSC - and is available on GitHub [https://github.com/GEMINI-multimorbidity/partialLDSC]. All analyses have been performed using version 0.1.0.

### Causal inference using Mendelian randomization

To better understand the causal relationship between BMI and each individual LTC, we performed two-sample Mendelian Randomization (MR) analyses. MR is a causal inference method that uses genetic variants as instrumental variables to estimate the causal effect of an exposure on an outcome. We used it to estimate the causal effect of intervening to lower BMI on each individual LTC (Supplementary Figure 2, Supplementary Note 2).

### Removing BMI causal effect from genetic correlations

For LTCs found to be strongly causally affected by BMI (23 LTCs, Supplementary Note 2), we used a recently proposed Bayesian GWAS approach to derive direct effect estimates [25,26]. The direct effects were estimated by taking out the causal effect of BMI from the observed association effect between each genetic variant and the condition. They correspond to the direct association between genetic variants and the condition, not via the BMI pathway, and can be seen as BMI-corrected GWAS summary statistics. We ran the analysis for each condition using the bGWAS R-package (https://github.com/nmounier/bGWAS - version 1.0.3) [25], selecting instruments for BMI using a p-value threshold of 5*10^-8^ with default values used for other parameters. We then used the summary statistics for the direct effects to re-estimate genetic correlations (denoted as bGWAS genetic correlations, 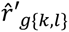) for 253 pairs. We only reported the results for the 246 pairs for which we detected a statistically significant difference between the unadjusted and the partial genetic correlation estimates. We then compared partial genetic correlations to bGWAS genetic correlations to understand if the difference between unadjusted and partial genetic correlations are likely to be driven by the causal effect of BMI on LTCs, or solely by the correlation between BMI and the LTCs, which could be due to other mechanisms.

### Estimating the causal effect of BMI on co-occurrence of LTCs and estimating the effect of intervening on BMI

To further understand the role of BMI in the co-occurrence of LTCs, we carried out additional analyses for the 15 pairs with the strongest difference, and no evidence of genetic correlation, after adjusting for BMI. First, we performed an additional GWAS in UK Biobank, defining cases for our analyses as individuals having been diagnosed with **both** LTCs (Supplementary Note 3). These GWAS summary statistics were then used to estimate the causal effect of BMI on the co-occurrence of each pair, using the same MR approach as for individual LTCs (Supplementary Note 2). The genetic associations used for MR are in standard deviation units for BMI (https://gwas.mrcieu.ac.uk/datasets/ieu-a-835/, 1SD = 4.77), and are log odds ratios for the outcomes. Therefore, for each pair *p*, the causal effect estimate 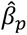 represents log odds ratios per 1 standard deviation (σ) increase in the BMI on the pair, compared to the actual observed BMI in the sample 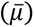:

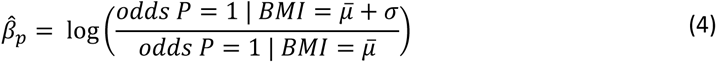

This causal effect estimate can then be used to evaluate how intervening to lower BMI would affect the number of people having both LTCs [27]. In practice, for each pair, we estimated the reduction in 1000 individuals 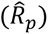 of reducing BMI by 1 SD:

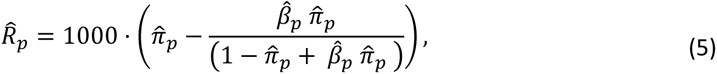

where 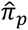 is the observed prevalence for pair *p*, using this risk reduction as the basis for further calculations. We used it to derive the number of cases that could be prevented from each pair following a hypothetical intervention in the sample used for our genetic analyses and estimated the impact on the prevalence of such an intervention.

## Results

### BMI plays an important role in explaining genetic correlations between common long-term conditions

From the 2485 pairs defined from the 71 LTCs, we detected a statistically significant difference when adjusting for BMI genetics for 1362 pairs (Table 1, Supplementary Table 2). These pairs encompassed 64 out of 71 LTCs. For most of these pairs (1078/1362), the partial genetic correlation estimates were weaker than the unadjusted ones, reflecting an attenuation of the genetic correlation when adjusting for BMI genetics.

**Table 1:**
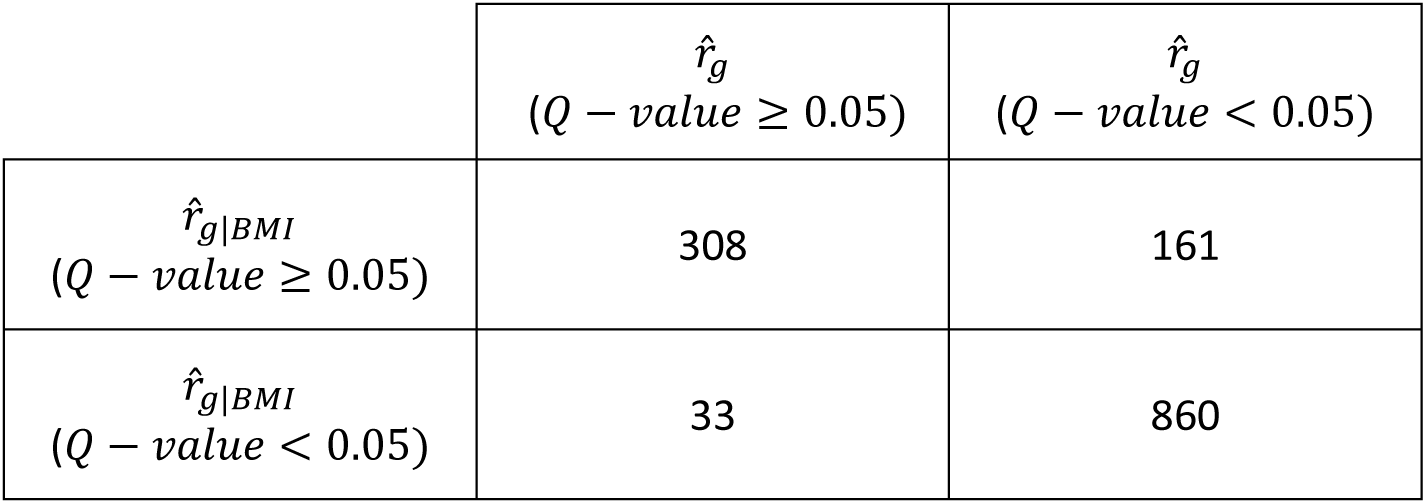
Description of the 1362 pairs with statistically significant differences (Q − value < 0.05 for the test statistics described in Equation 4), according to the statistical significance of unadjusted 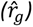 and partial 29 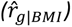 genetic correlation estimates.

| | $\hat{r}_g$<br>($Q - value \geq 0.05$) | $\hat{r}_g$<br>($Q - value < 0.05$) |
| --- | --- | --- |
| $\hat{r}_{g\vert BMI}$<br>($Q - value \geq 0.05$) | 308 | 161 |
| $\hat{r}_{g\vert BMI}$<br>($Q - value < 0.05$) | 33 | 860 |

In most of the pairs (860/1362) for which a statistically significant difference was observed, we found evidence of shared genetics both before and after adjusting for BMI, indicating that while BMI influences the genetic correlation for these pairs, it does not account for all of it. These include 131 *within-domain* pairs, and 769 *cross-domain* pairs, many of them encompassing LTCs from the “diseases of the circulatory system”, but also from the “diseases of the skin and subcutaneous tissue” and the “diseases of the digestive system” domains. For most of these pairs, the partial genetic correlation estimates were smaller (740/860). The majority of the 120 pairs where the partial genetic correlation estimates were larger were related to anxiety disorder (30), osteoporosis (27), schizophrenia and delusional disorders (12), as well as tinnitus (11). The unadjusted and the partial genetic correlation estimates for the 20 pairs of these 860 with the strongest difference are presented in **Error! Reference source not found.**. Amongst these 20 top pairs, 12 distinct LTCs are represented, with some conditions common between pairs with cholelithiasis in 6 pairs and carpal tunnel syndrome, gout and chronic kidney disease in 7 pairs. This illustrates that BMI genetics tends to explain a larger proportion of genetic correlation in pairs that contain specific LTCs. Furthermore, these LTCs are also amongst the ones with the strongest genetic correlations with BMI (Supplementary Table 1), and most strongly causally affected by BMI in our MR results (Supplementary Note 2). For 471 of these 860 pairs, our MR analysis showed that BMI is likely causal for both LTCs, providing further evidence that BMI is the likely source of the genetic correlation.

For more than 10% of the pairs with a significant difference (161/1362), we showed that BMI is likely to account for a substantial component of the genetic correlation. For these pairs, BMI adjustment resulted in a greatly reduced partial genetic correlation estimate consistent with no remaining genetic similarity between the two LTCs. Many of these pairs (76/161) included at least one condition from the “*diseases of the circulatory system*” domain, and almost all the pairs were *cross-domain* pairs (154/161)for example 24 pairs had one condition from the “*diseases of the circulatory system*” domain, and one condition from the “*diseases of the musculoskeletal system and connective tissue*” domain.

For instance, we observed complete attenuation of the genetic correlation between: type 2 diabetes and osteoarthritis 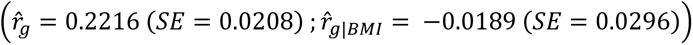; gout and osteoarthritis 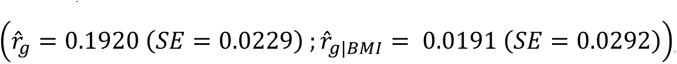; and gout and sleep apnoea 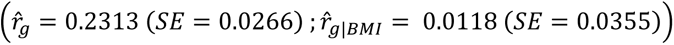 (Supplementary Table 2, Figure 2). For 93 of these pairs, including all the ones in Figure 2, our MR analysis detected a statistically significant causal effect of BMI on both LTCs.

**Figure 1.**
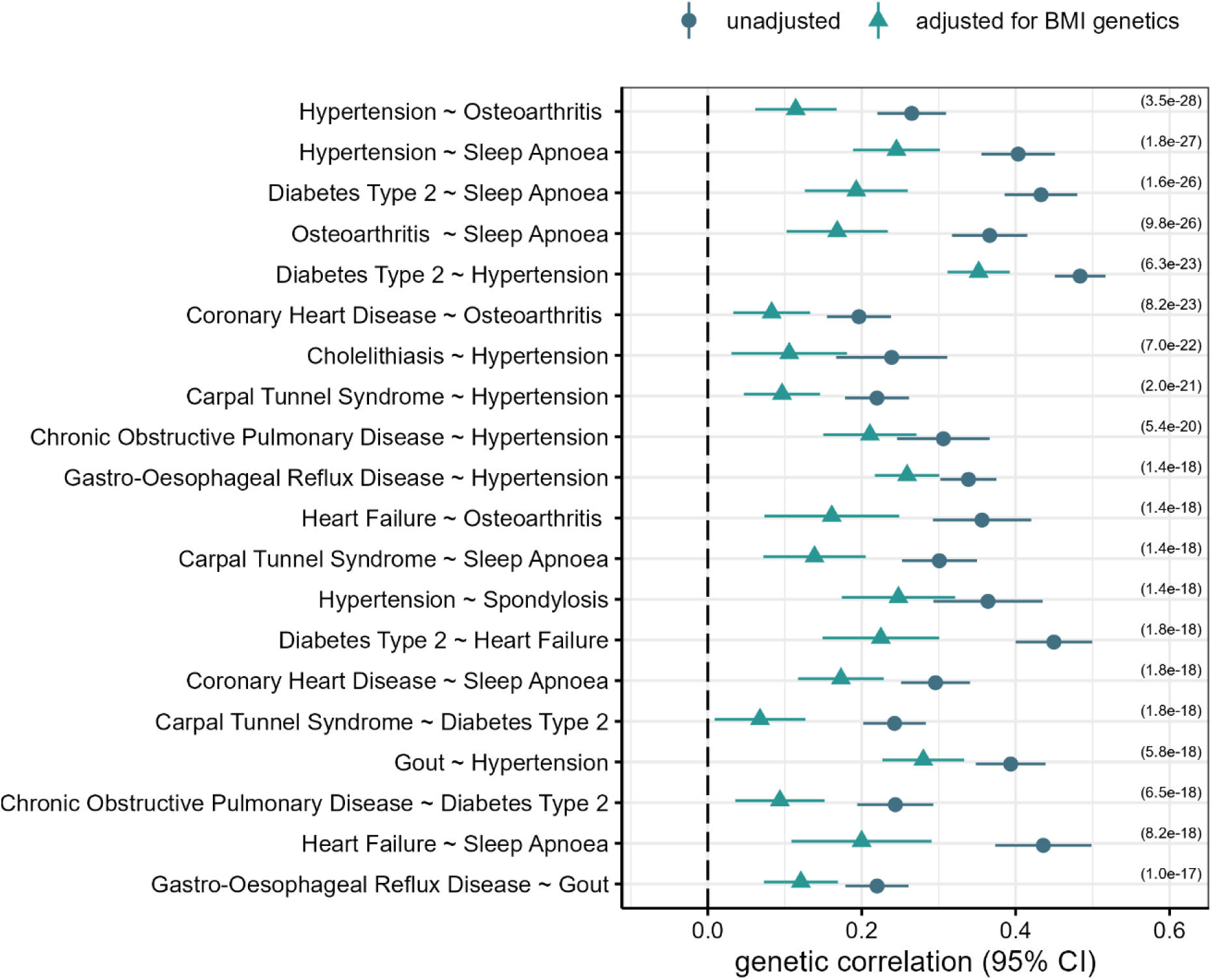
Unadjusted (blue circle) and partial (blue-green triangle) genetic correlation estimates and 95% confidence intervals for the 20 pairs with the strongest difference (Q − value indicated for each pair on the right) and evidence of shared genetics both before and after adjusting for BMI.

**Figure 2.**
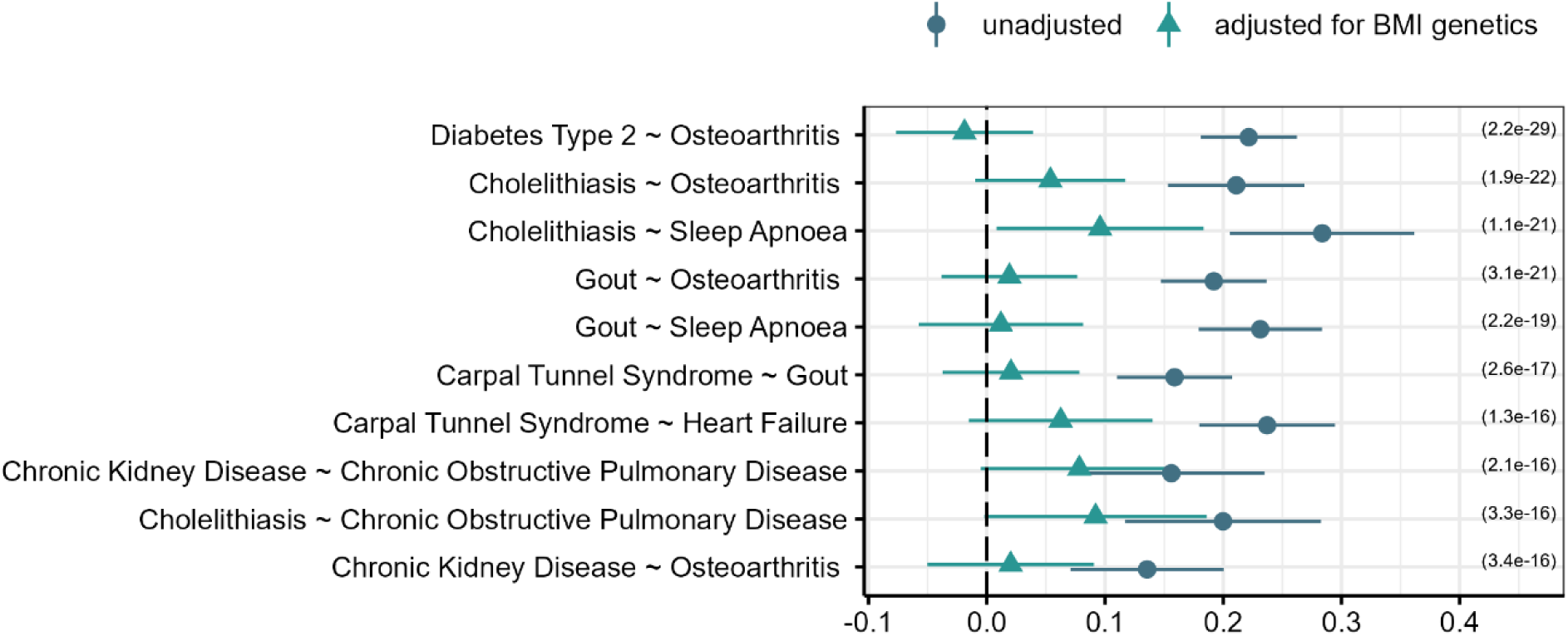
Unadjusted (blue circle) and partial (blue-green triangle) genetic correlation estimates and 95% confidence intervals for the 10 pairs with the strongest difference (Q − value indicated for each pair on the right) and evidence of shared genetics only before adjusting for BMI.

For 33 LTC pairs, there was no evidence (at FDR <0.05) of genetic correlation, but on adjustment for the genetics of BMI, a residual genetic effect was observed (Supplementary Table 2, *Figure 3*). These results are consistent with BMI masking shared genetic mechanisms between the two LTCs. Most of these pairs were cross-domain pairs (28/33) and notably, 14 of these pairs were related to osteoporosis. This can be explained by the fact that lower BMI has been shown to be associated with higher osteoporosis risk [28]whereas for most other LTCs, higher BMI is suspected to be risk increasing. This is further supported by our MR analysis, as we found a statistically significant causal effect of lower BMI on osteoporosis (Supplementary Note 2). For 19 of these pairs, our MR results showed that BMI is likely causal for both LTCs, but protective for one of them, consistent with the partial genetic correlation being stronger than the unadjusted genetic correlation.

**Figure 3.**
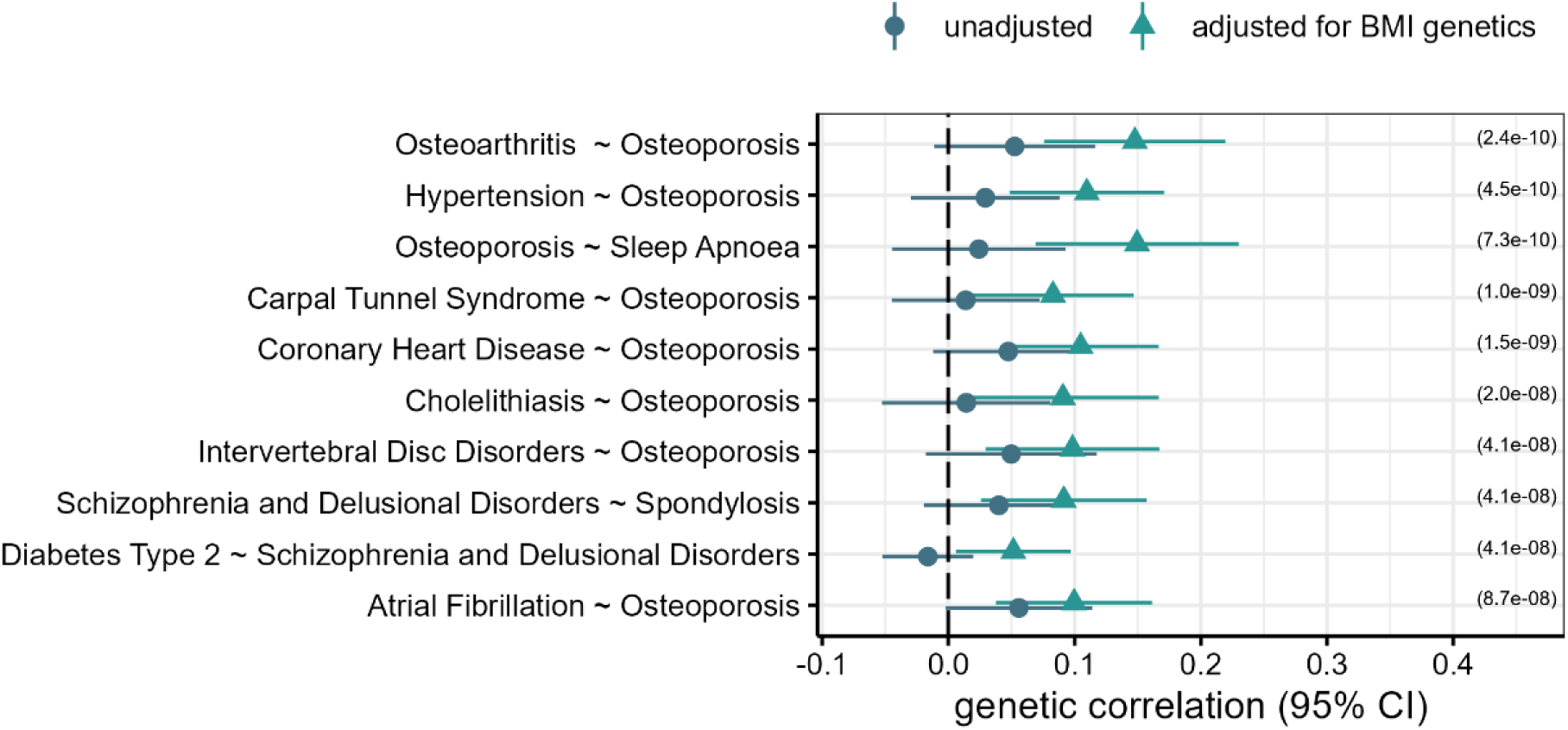
Unadjusted (blue circle) and partial (blue-green triangle) genetic correlation estimates and 95% confidence intervals for the 10 pairs with the strongest difference (Q − value indicated for each pair on the right) and evidence of shared genetics only after adjusting for BMI.

For 1123 LTC pairs, adjusting for BMI genetics had no statistically significant effect on the genetic correlation between them, suggesting that other mechanisms are giving rise to the genetic similarity between these LTCs. For example, the strong genetic correlation between allergic rhinitis and bursitis 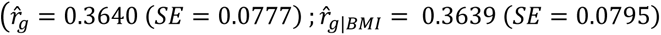; difference *Q* − *value* = 0.98), or between anxiety and schizophrenia 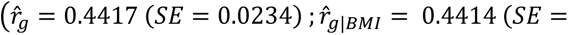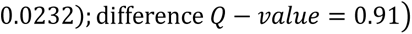, remain similar after adjusting for BMI genetics.

These results were consistent with those obtained using less recent BMI data, excluding UK Biobank, with partial correlation estimates that were highly similar (correlation = 0.9974, Supplementary Figure 3-A), with evidence of the analysis using the more recent data being more powered (Supplementary Table 3, Supplementary Table 4, Supplementary Figure 3-B).

### BMI acts as a common risk factor in multimorbidity

To compare our partial genetic correlations to another method (bGWAS) we estimated pairwise genetic correlations accounting for the estimated causal effect of BMI on *both* LTCs, for 246 pairs. We studied the 23 LTCs that were strongly causally affected by BMI and a subset of LTC pairs for which a statistically significant difference between unadjusted and partial genetic correlation estimates were observed. We used bGWAS to take out the causal effect of BMI on each individual LTC and re-estimated pairwise genetic correlations. These differ from the partial correlation estimates since they only remove the BMI genetics estimated to be due to the causal role of BMI on each condition. We observed a strong agreement between the partial genetic correlation estimates and the bGWAS correlation estimates, with some of the bGWAS genetic correlation estimates being stronger (less attenuated) than the partial ones (Supplementary **Error! Reference source not found.**-A, Supplementary Table 5), suggesting that partial genetic correlation estimates might capture additional associations between BMI and the LTCs that are not due to these causal relationships. See Supplementary Note 4 for details.

For a subset of pairs, the 15 pairs with the strongest difference and no evidence of genetic correlation after adjusting for BMI genetics, we performed genetic analysis for each pair and further investigated the role of BMI using MR results. We showed that higher BMI had a strong risk-increasing causal effect on every condition within these pairs. In addition, the unadjusted genetic correlation estimates were all positive, and higher BMI was shown to be risk increasing for all LTCs (Supplementary Table 6). This suggests that the attenuation of the genetic correlation is likely due to BMI acting as common risk factor. This is further confirmed by the MR analysis on the pairs, with all causal effect estimates being positive and statistically significant (*p* − *value* < 0.05/64, Bonferroni correction) (Supplementary Table 6).

For these pairs with directionally consistent individual causal effects, we also estimated how intervening on BMI would affect the number of people having **both** LTCs. To do so, we calculated the reduction in the number of cases for 1000 individuals expected by a one SD BMI reduction. Results were highly correlated with the observed prevalence of the pairs in our sample and lowering BMI had a stronger effect on pairs with higher prevalence (Supplementary Table 6). Results for the 5 pairs with a prevalence above 1% are presented in Table 2. For example, 16 out of 1000 people having both chronic kidney disease and osteoarthritis, and 9 out of 1000 people having both type 2 diabetes and osteoarthritis, would not have both LTCs after a one SD BMI intervention (1SD = 4.77). In the UK Biobank sample used for our analysis, this would translate into a 4.6% reduction in prevalence for the co-occurrence of chronic kidney disease and osteoarthritis, and 3.3% reduction in prevalence for the co-occurrence of type 2 diabetes and osteoarthritis. We postulate that the impact of such an intervention in the general population is likely to be stronger, since the prevalence of these pairs of LTC in UK Biobank is lower than in the general population (Supplementary Table 6), due to healthy volunteer bias in UK Biobank [29].

**Table 2.** Mendelian randomization and BMI intervention results for 5 pairs with a prevalence above 1% in our sample. For each pair of LTC (condition 1 and condition 2), the unadjusted genetic correlation estimate and standard error 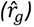, the partial genetic correlation estimate and standard error 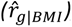, as well as the causal effect estimate 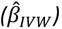, its standard error and the corresponding p-value, of BMI on the pair, and the reduction in the number of cases with both LTCs when reducing BMI by one SD for 1000 individuals 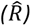, and the prevalence of cases with both LTCs in percentage in UK Biobank before (π UK Biobank) and after an hypothetical intervention (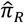 UK Biobank), are reported.

| condition 1 | condition 2 | $\hat{r}_g$<br>(SE) | $\hat{r}_{g\vert BMI}$<br>(SE) | $\hat{\beta}_{IVW}$<br>(SE ; $p - value$) | $\hat{R}$ | $\pi$<br>UK<br>Biobank | $\hat{\pi}_R$<br>UK<br>Biobank |
| --- | --- | --- | --- | --- | --- | --- | --- |
| Chronic Kidney Disease | Osteoarthritis | 0.1356<br>(0.0330) | 0.0202<br>(0.0359) | 0.38<br>(0.0835 ; 4.2e-06) | 16.76 | 2.752 | 2.706 |
| Diabetes Type 2 | Osteoarthritis | 0.2216<br>(0.0208) | -0.0189<br>(0.0296) | 0.71<br>(0.1820 ; 9.4e-05) | 9.69 | 3.434 | 3.401 |
| Cholelithiasis | Osteoarthritis | 0.2110<br>(0.0295) | 0.0538<br>(0.0323) | 0.63<br>(0.0775 ; 6.0e-16) | 8.83 | 2.406 | 2.385 |
| Asthma | Chronic Kidney Disease | 0.0829<br>(0.0353) | 0.0158<br>(0.0361) | 0.35<br>(0.1132 ; 2.1e-03) | 7.10 | 1.092 | 1.085 |
| Gout | Osteoarthritis | 0.1920<br>(0.0229) | 0.0191<br>(0.0292) | 0.61<br>(0.0963 ; 3.2e-10) | 6.47 | 1.657 | 1.646 |

## Discussion

Clinical trials and Mendelian randomization studies have shown that obesity is very likely to cause many individual LTCs [30]. However, the role of higher BMI in the co-occurrence of two LTCs and the extent to which it accounts for some, or all of the co-occurrence is less certain. In this study we combined genetics data from very large studies with a novel statistical analysis technique to estimate the extent to which higher BMI influences the genetic similarity between pairs of LTCs. Our analyses indicated that for a large proportion of pairs of LTCs, higher BMI is likely to explain part of the genetic correlation, but that other factors are likely to also contribute. For 55% of LTC-pairs tested we observed a change in the genetic correlations when accounting for BMI genetics. Using causal inference methods, we confirmed that BMI acts as a common risk factor for a subset of these pairs, and that intervening on BMI could help reduce the prevalence of pairs of multimorbid LTCs.

Our analyses indicated that for 36% (740/2485) of LTCs pairs we studied, higher BMI could explain a significant proportion of their genetic correlation. Of these, 161 pairs were identified for which the estimates were consistent with a null partial genetic correlation, suggesting that the genetic similarity between these pairs of LTCs is *entirely* explained by the BMI. These included many pairs that spanned traditional disease domains, especially those involving osteoarthritis and metabolic diseases, and COPD and metabolic diseases. For some of these pairs, previous studies have observed a similar strong attenuating effect of BMI, such as for type 2 diabetes and osteoarthritis and gout and sleep apnoea [31,32].

Thirty-three pairs (about 1%) of LTCs showed no evidence of unadjusted genetic similarity, but after accounting for the genetic effect of BMI, a partial genetic correlation emerged. These results suggest that shared genetic causes may exist for some pairs, but they could be masked by BMI having effects in opposite directions. Most of these pairs were related to osteoporosis, which may be explained by the fact that the direction of the effect of BMI on osteoporosis is opposite to the one on most other LTCs [33]. We observed for example a positive genetic correlation between osteoporosis and stroke, only after accounting for BMI genetics. Because genetic risk usually reflects life-long exposure, this correlation suggests the presence of shared risk factors for osteoporosis and metabolic traits, although we cannot rule out a genetic correlation resulting from reduced mobility and bone load reduction in stroke patients [34].

Our results corroborate the role of BMI acting as a common risk factor for various pairs of LTCs. We have used causal inference approaches to test the causal relationship between BMI and the different LTCs, and we used the bGWAS approach to re-estimate genetic correlation estimates, adjusting only for the part of the genetic correlation that is driven by the causal effect of BMI on the LTCs. Overall, we observed a strong agreement between partial and bGWAS genetic correlation estimates, providing further evidence that BMI does act as common risk factor. However, the bGWAS genetic correlation estimates were in general stronger, suggesting that the correlation between BMI and the LTCs is not entirely explained by the causal effect and that more complex relationships may exist. In addition, the effects derived from bGWAS would only capture linear causal effects, so any non-linear effects would not be accounted for, possibly explaining why the bGWAS genetic correlations estimates were stronger. We also showed that BMI has a strong, risk-increasing, causal effect on some pairs of LTCs, and that lowering BMI could help reduce the prevalence of these pairs.

Many of the LTC pairs include those from across traditional clinical domains, and for a majority of these pairs, the attenuation after accounting for BMI was as strong as that for within-domain pairs. For example, based on our genetic approach, the role of BMI in explaining the co-occurrence of COPD and several metabolic LTCs and osteoarthritis and several metabolic LTCs, is just as strong as the role of BMI in explaining the co-occurrence of two metabolic LTCs. The reasons for these findings require further investigation but our results suggest that interventions aimed at weight loss should monitor these additional LTCs more closely, or conversely, patients with a combination of obesity and a metabolic condition should be monitored for less obvious non-metabolic LTCs.

The definition of obesity, and other obesity-related measures and phenotypes that can be used as proxies in public health research, is critical [35] and which one is the best predictor for disease risk might depend on the condition [36–42]. For that reason, we believe that results using BMI, as a continuous phenotype that can be easily measured in the general population, are less likely to be biased and are easier to interpret. Further analyses, to better understand how exactly BMI affects each condition could be performed, using clustering-based MR approaches for instance [43]. Other obesity-related phenotypes should be used to investigate the effect of body fat distribution, like waist-to-hip ratio for example, and these analyses would benefit from a sex-specific approach, to better account for the known sex-specific genetic architecture of such phenotypes [44,45].

This work focused on individuals of European descent, mostly because of data availability, and analyses are needed in other ethnic groups, as it is known that both BMI distribution [46] and its effect on LTCs may vary depending on ethnicity [47,48], to further understand the role of BMI in multimorbidity in these groups. It is important to note that the relative paucity of data from people of non-European ancestry has a disproportionately large effect on the utility of genetics for multimorbidity, because missing data from only one condition will affect multiple pairs.

This work not only has important implications for research that aims at identifying shared biological pathways between pairs of LTCs but also has direct implications for potential intervention. We showed that intervening on BMI would directly impact the prevalence of pairs of LTCs, such as type 2 diabetes and chronic kidney disease, or type 2 diabetes and osteoarthritis. This is particularly important with the recent commercialisation of weight loss drugs that could be used to reduce the co-occurrence of LTCs through a better weight management strategy. This work focused on individuals of European descent, mostly because of data availability, and analyses are needed in other ethnic groups, as it is known that both BMI distribution [46] and its effect on LTCs may vary depending on ethnicity [47,48], to further understand the role of BMI in multimorbidity in these groups.

## Data Availability

All data produced in the present work are contained in the manuscript

## Funding

This work was supported by the UK Medical Research Council [grant number MR/W014548/1]. This study was supported by the National Institute for Health and Care Research (NIHR) Exeter Biomedical Research Centre (BRC), the NIHR Leicester BRC, the NIHR Oxford BRC, the NIHR Peninsula Applied Research Collaboration, and the NIHR Health Tech Research Centre. JM is funded by an NIHR Advanced Fellowship (NIHR302270). The views expressed are those of the authors and not necessarily those of the NIHR or the Department of Health and Social Care.

## Acknowledgements

We acknowledge all members of the GEMINI Consortium. This includes researchers Olivia Murrin, Carlos Gallego-Moll, Albert Roso-Llorach, Lucía A Carrasco-Ribelles, Chris Fox, Louise M Allan, Xiaoran Liang, Ruby M Woodward, Deniz Türkmen, Kate Boddy, Jose M Valderas, Sara M Khalid, Sally E Lamb, David Melzer, and Concepción Violán, in addition to patient representatives Mary Mancini and Leon Farmer. The whole GEMINI team were central to developing the LTC definitions, observational associations in population representative primary care cohorts, genetic associations between LTC pairs, and developing and refining the research question(s) and results.

This research has been conducted using the UK Biobank Resource under Application Number 9072. We want to acknowledge the participants and investigators of the FinnGen study. We also thank the authors and study participants for the published GWAS consortium meta-analyses utilized in this report (see [11] and the project GitHub pages [https://github.com/GEMINI-multimorbidity] for details and citation). The authors would like to acknowledge the use of the University of Exeter High-Performance Computing (HPC) facility in carrying out this work.

## Disclosures

JB is a part time employee of Novo Nordisk Research Centre Oxford, limited. TF has consulted for several pharmaceutical companies. All other authors have no disclosures to declare.

## Contributions

Conceptualization: NM, LP, FD, TF, JB. Data Curation NM, BV, LP: Formal Analysis: NM, BV, LP. Funding Acquisition JM, JD, FD, LP, JB, TF. Interpretation: NM, BV, JM, JD, FD, LP, TF, JB. Investigation: NM, JB. Resources: NM, BV, LP. Software: NM Supervision: JB, TF, LP, FD. Writing – Original Draft: Writing – NM, LP, TF, JB. Review & Editing: NM, BV, JM, JD, FD, LP, TF, JB.

## Notes

### Competing Interest Statement

Jack Bowden is a part time employee of Novo Nordisk Research Centre, Oxford limited. Tim Frayling has consulted for several pharmaceutical companies. All other authors have no competing interests.

### Author Declarations

All data used in this research is publicly available to the research community.

