## supplementary methods and figures for "Genetics identifies obesity as a shared risk factor for co-occurring multiple long-term conditions"

*Mounier et al. 2024*

#### Supplementary Notes

**Supplementary Note 1 – Block Jackknife**

**Supplementary Note 2 – Mendelian Randomization**
**Supplementary Note 3 – GWAS of pairs**
**Supplementary Note 4 – comparison to bGWAS**

#### Supplementary Tables

**Supplementary Table 1 (available in STable1.xlsx)**
**Supplementary Table 2 (available in STable2.xlsx)**
**Supplementary Table 3**
**Supplementary Table 4 (available in STable4.xlsx)**
**Supplementary Table 5 (available in STable5.xlsx)**
**Supplementary Table 6 (available in STable6.xlsx)**
**Supplementary Table 7 (available in STable7.xlsx)**

#### Supplementary Figures

**Supplementary Figure 1**
**Supplementary Figure 2**
**Supplementary Figure 3**
**Supplementary Figure 4**
**Supplementary Figure 5**

### Supplementary Note 1 – Block Jackknife

Jackknife resampling is a technique, closely related to bootstrapping, that is often used for bias and variance estimation. A jackknife estimator is derived by creating subsamples, omitting one observation, and aggregating the parameters estimates from each subsample [1]. In statistical genetics, when using SNP-trait associations, the standard jackknife technique can not readily be used because of the underlying LD structure and the correlation between adjacent SNPs. Therefore, it has been proposed to omit blocks of observations rather than single observations, and several approaches have been using block jackknife for variance estimation [2–6] or to mitigate biases [7].

In practice, we implemented the following block jackknife procedure, defining $J$ jackknife blocks, to estimate the variance of each element of the genetic covariance matrix, $\rho_g$. Let's consider $\hat{\rho}_g$ the estimator obtained from the entire sample. For each block $j$, we created the $j$-th jackknife subsample by removing the $j$-th block from the GWAS summary statistics. We used this subsample to estimate the leave-one-out value $\left(\hat{\rho}_g{}^j\right)$, and derive the pseudo value $\left(\tilde{\rho}_g{}^j\right)$, when omitting block $j$ : $\tilde{\rho}_g{}^j = J\,\hat{\rho}_g - (J-1)\,\hat{\rho}_g{}^j$ . The variance of the pseudo values $(\tilde{s}^2)$ can then be used to obtain an estimate of the variance of the genetic covariance matrix estimate: $Var\left(\hat{\mathrm{P}}\right) = \frac{\tilde{s}^2}{J}$.
Similarly, we defined the partial genetic covariance matrix ($\hat{\rho}_{g\,|\,x}$), the genetic correlation matrix $\left(\hat{r}_g\right)$ and the partial genetic correlation matrix $\left(\hat{r}_{g\,|\,x}\right)$ (as described in the *Partial genetic correlation* subsection) and derived leave-one-out and pseudo-values to estimate their variance.

While using blocks helps reduce the correlation between the observations that are omitted and the observations that are not in each subsample, it is unclear what the ideal number of blocks is. Previous implementations of LD-score regression have been using a default value of 200 blocks [2,3], or advising to increase the number of blocks when the number of elements in the covariance matrix increases [4]. In our analyses, we investigated pairwise genetic correlations between 71 conditions, hence estimating almost 2,500 parameters. However, when using a larger number of blocks, the size of each block decreases and smaller blocks are more susceptible to only contain SNPs that are correlated with SNPs in adjacent blocks, hence increasing the between-blocks correlation and defeating the purpose of using blocks. We assessed how the number of blocks used influenced our results, varying the number of blocks used from 200 to 4500 (Supplementary Figure 4). We showed that as the number of blocks increases, the number of pairs reaching significance (for both unadjusted, partial genetic correlations, as well as for the difference between the two) increases. This is because the variance estimates are getting smaller and could be due to the correlation between blocks getting larger and the number of blocks $J$ no longer reflecting the number of independent observations. For this reason, we decided to use 200 blocks for our analyses.

### Supplementary Note 2 – Mendelian Randomization

Mendelian Randomization (MR) MR is a causal inference method that uses genetic variants as instrumental variables to estimate the causal effect of an exposure on an outcome (Supplementary Figure 2). We used the inverse-variance weighed estimator, implemented in the `TwoSampleMR` R-package (version 0.5.7) [8] to estimate the causal effect of BMI on each of the LTC, denoted as $\hat{\beta}_{IVW}$. For these analyses, we used BMI GWAS summary statistics that do not include UK Biobank [9] and use the default parameters to select strong independent instruments (up to 68 used). We limited these analyses to those conditions that were in at least one pair of LTCs for which adjusting for the genetic of BMI resulted in a change in genetic correlation (64 conditions, from the 1362 pairs identified comparing unadjusted and partial genetic correlations).

We detected a significant causal effect of BMI on 41 conditions ($p - value < 0.05$), with 23 of them surviving multiple testing correction ($p - value < 0.05/64$, Bonferroni correction) (Supplementary Table 7). The ten conditions the most strongly causally affected by BMI are type 2 diabetes $\left(\hat{\beta}_{IVW} = 0.8288, \mathrm{SE} = 0.0679, \mathrm{p} = 2.6 \cdot 10\mathrm{e}^{-34}\right)$, cholelithiasis $\left(\hat{\beta}_{IVW} = 0.4858, \mathrm{SE} = 0.0435, \mathrm{p} = 6 \cdot 10\mathrm{e}^{-29}\right)$, sleep apnoea $\left(\hat{\beta}_{IVW} = 0.5951, \mathrm{SE} = 0.0589, \mathrm{p} = 6 \cdot 10\mathrm{e}^{-24}\right)$, heart failure $\left(\hat{\beta}_{IVW} = 0.5154, \mathrm{SE} = 0.0529, \mathrm{p} = 2 \cdot 10\mathrm{e}^{-22}\right)$, gout $\left(\hat{\beta}_{IVW} = 0.3750, \mathrm{SE} = 0.0408, \mathrm{p} = 4 \cdot 10\mathrm{e}^{-20}\right)$, osteoarthritis $\left(\hat{\beta}_{IVW} = 0.3619, \mathrm{SE} = 0.0397, \mathrm{p} = 7 \cdot 10\mathrm{e}^{-20}\right)$, thromboembolic diseases $\left(\hat{\beta}_{IVW} = 0.4701, \mathrm{SE} = 0.0537, \mathrm{p} = 2 \cdot 10\mathrm{e}^{-18}\right)$, hypertension $\left(\hat{\beta}_{IVW} = 0.4340, \mathrm{SE} = 0.0539, \mathrm{p} = 8 \cdot 10\mathrm{e}^{-16}\right)$, atrial fibrillation $\left(\hat{\beta}_{IVW} = 0.3911, \mathrm{SE} = 0.0492, \mathrm{p} = 2 \cdot 10\mathrm{e}^{-15}\right)$, carpal tunnel syndrome $\left(\hat{\beta}_{IVW} = 0.4145, \mathrm{SE} = 0.0593, \mathrm{p} = 3 \cdot 10\mathrm{e}^{-12}\right)$ and coronary heart disease $\left(\hat{\beta}_{IVW} = 0.2955, \mathrm{SE} = 0.0450, \mathrm{p} = 5 \cdot 10\mathrm{e}^{-11}\right)$. BMI had a significant negative causal effect on seven conditions, the two being the most strongly causally affected being breast cancer $\left(\hat{\beta}_{IVW} = -0.2849, \mathrm{SE} = 0.0536, \mathrm{p} = 1 \cdot 10\mathrm{e}^{-7}\right)$ and osteoporosis $\left(\hat{\beta}_{IVW} = -0.1971, \mathrm{SE} = 0.0628\ \mathrm{p} = 2 \cdot 10\mathrm{e}^{-3}\right)$.

Similarly, we used the same approach (same BMI data and same parameters) to estimate the causal effect of BMI on specific pairs of LTCs and observed a statistically significant causal effect on all the pairs after adjusting for multiple testing ($p - value < 0.05/15$, Bonferroni correction) (Supplementary Table 6 (available in STable6.xlsx)).

#### Supplementary Note 3 – GWAS of pairs

For a subset of pairs, the 15 pairs with the strongest difference and no evidence of genetic correlation after adjusting for BMI (Supplementary Table 6 (available in STable6.xlsx)), we performed genome-wide association studies (GWAS).

To perform the genetic analyses using pairs of LTCs, we used the UK Biobank, a large population-based prospective study [10] and restricted our analyses to 450,197 individuals of European ancestry (identified using genomic principal components); 205,737 males and 244,260 females. We used both primary-care linked data (available for 209,387 participants, censoring date: 28/02/2016 – Read v2 and CTV3 codes, truncated to 5 bytes) and hospital diagnoses (available for all participants, censoring date: 31/10/2022 - ICD-10 codes) to identify cases for each condition, using the definitions described in [11].

We then focussed on 15 pairs of conditions, defining cases for our analyses as individuals having been diagnosed with both conditions, and controls as individuals having been diagnosed with only one of the conditions, or none of them. The number of cases for each pair of condition in our sample can be found in Supplementary Table 6. We performed GWAS for the 15 pairs using REGENIE (v3.1.3) [12], adjusting for the covariates: age at baseline, sex, genotyping chip, and assessment centre. We then implemented a quality-control step, restricting the included variants to those with a minor allele frequency (MAF) of > 0.1%, and an imputation INFO score ≥0.3.

#### Supplementary Note 4 – Comparison to bGWAS

To better understand if the differences between unadjusted and partial genetic correlations estimates could be explained by a causal effect of BMI on both conditions, we estimated pairwise genetic correlations, accounted for the estimated causal effect of BMI on both conditions, for 246 pairs. We used a subset of pairs derived from the 23 conditions that were strongly causally affected by BMI, and for which a statistically significant difference between unadjusted and partial genetic correlation estimates was observed. We used bGWAS [13] to take out the causal effect of BMI on each individual LTC and re-estimated pairwise genetic correlations. The genetic correlation estimates obtained using bGWAS differ from the partial correlation estimates since they do not take out the effect of BMI genetics in general, but only the part that is estimated to be due to the causal role of BMI on each condition. For these pairs, we observed a strong agreement between the partial genetic correlation estimates and the bGWAS correlation estimates, with some of the bGWAS genetic correlation estimates being stronger than the partial ones (Supplementary

-A, Supplementary Table 5). This suggests that adjusting for BMI genetics is to some extent similar to adjusting for the causal effect of BMI, but that partial genetic correlation estimates might capture additional associations between BMI and the conditions that are not due to these causal relationships. Overall, all the bGWAS genetic correlation estimates were much more consistent with the partial ones than with the unadjusted ones (Supplementary Table 5, Supplementary

5-B), and most of the statistically significant differences observed between the unadjusted and the partial genetic correlation estimates for these pairs of LTCs are likely to be driven by a causal effect of BMI on both conditions, corroborating its role as a common risk factor. The partial and the bGWAS genetic correlation estimates were similar to each other and lower than the unadjusted ones for many pairs, including hypertension and hypothyroidism $\left(\hat{r}_g = 0.1519\ (SE = 0.328);\ \hat{r}_{g|BMI} = 0.1005\ (SE = 0.0318)\ ;\hat{r}'_g = 0.1046\ (SE = 0.0358)\right)$ or atrial fibrillation and thromboembolic diseases $\left(\hat{r}_g = 0.1547\ (SE = 0.329);\ \hat{r}_{g|BMI} = 0.0898\ (SE = 0.0359)\ ;\hat{r}'_g = 0.0884\ (SE = 0.0352)\right)$. For example, while the unadjusted genetic correlation between gout and osteoarthritis was strong $\left(\hat{r}_g = 0.1920\ (SE = 0.0229)\right)$, both the partial and the bGWAS genetic correlation estimates were consistent with an absence of genetic association after adjusting for BMI $\left(\hat{r}_{g|BMI} = 0.0191\ (SE = 0.0292)\ ;\hat{r}'_g = 0.0568\ (SE = 0.0261)\right)$. For some pairs, the bGWAS genetic correlation estimates were smaller than the unadjusted ones, but larger than the partial ones, as seen for heart failure and sleep apnoea $\left(\hat{r}_g = 0.4360\ (SE = 0.319);\ \hat{r}_{g|BMI} = 0.1999\ (SE = 0.0465)\ ;\hat{r}'_g = 0.2465\ (SE = 0.0410)\right)$ for instance.

*<u>Supplementary Table 1</u> (available in STable1.xlsx)*
*Description of the 71 long term conditions used.*

*<u>Supplementary Table 2</u> (available in STable2.xlsx)*
*Partial genetic correlation results, adjusting for BMI genetics (data from 2018).*

*<u>Supplementary Table 3</u>*
*Description of the 1199 pairs with statistically significant differences ($Q - value < 0.05$), according to the statistical significance of unadjusted and partial genetic correlation estimates, using BMI data from 2015. Numbers between parentheses correspond to the number of pairs in common with the main analysis using BMI data from 2018 in green, the number of pairs unique to this analysis in blue, and the number of pairs unique to the main analysis in black.*

| | $\hat{r}_g$ ($Q - value \geq 0.05$) | $\hat{r}_g$ ($Q - value < 0.05$) |
|---|---|---|
| $\hat{r}_{g\|BMI}$ ($Q - value \geq 0.05$) | 285 (243 + 42 - 65) | 111 (108 + 3 - 53) |
| $\hat{r}_{g\|BMI}$ ($Q - value < 0.05$) | 28 (24 + 4 - 9) | 775 (717 + 58 - 143) |

*<u>Supplementary Table 4</u> (available in STable4.xlsx)*
*Partial genetic correlation results, adjusting for BMI genetics (data from 2015).*

*<u>Supplementary Table 5</u> (available in STable5.xlsx)*
*Direct genetic correlation results, using bGWAS to adjust for the causal effect of BMI on the conditions (data from 2015).*

*<u>Supplementary Table 6</u> (available in STable6.xlsx)*
*Genetic correlation, mendelian randomization and BMI intervention results for the 15 pairs with the strongest difference and no evidence of genetic correlation after adjusting for BMI.*

*<u>Supplementary Table 7</u> (available in STable7.xlsx)*
*Causal inference results: IVW-MR causal effect estimates of BMI on each individual LTC.*

*Supplementary Figure 1*

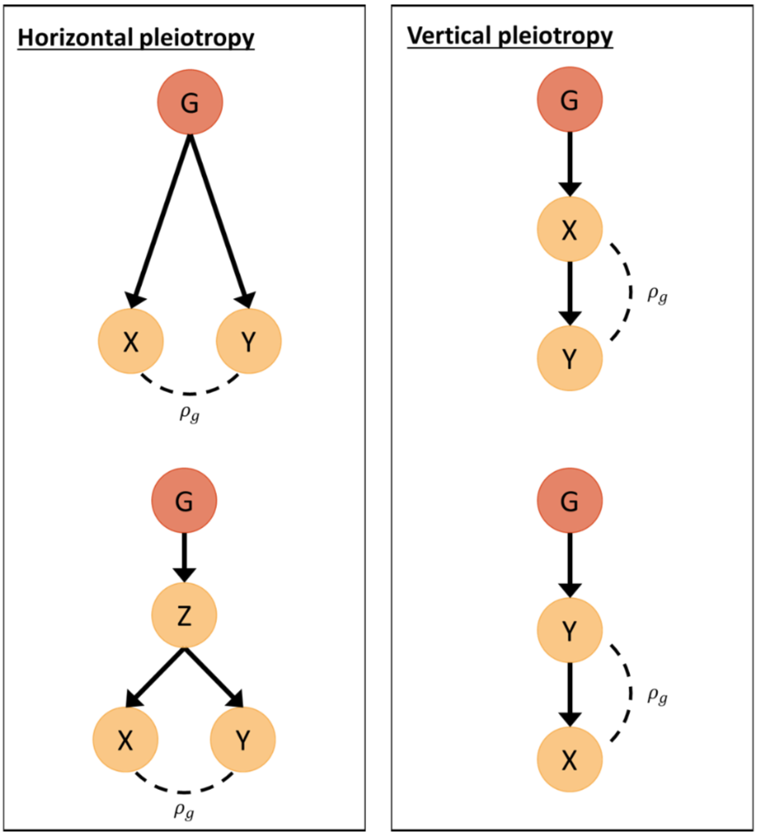


*Illustration of horizontal and vertical pleiotropy. In horizontal pleiotropy, the genetic variant (G) affects directly the risk of both diseases (X and Y) or indirectly through an intermediate phenotype (Z). In vertical pleiotropy, the genetic variant directly affects one of the diseases, and there is causal relationship between the two diseases. Both phenomena give rise to genetic covariance between the two diseases ($\rho_g$).*
*Figure adapted from* [14].

*Supplementary Figure 2*

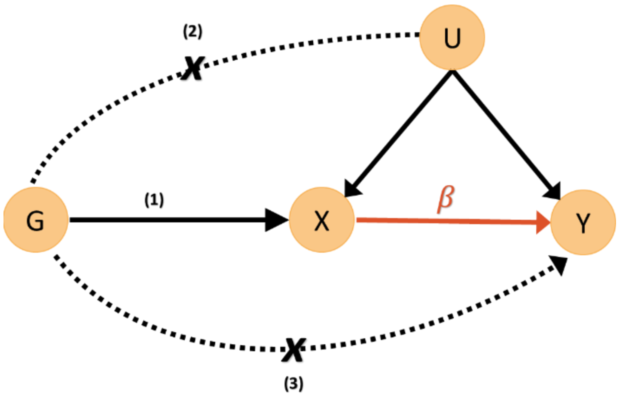


*Directed acyclic graph representing the MR model used to estimate the causal effect β of an exposure X on an outcome Y and its main assumptions: (1) Relevance – instrumental variables, denoted by G, are strongly associated with the exposure. (2) Exchangeability – G is not associated with any confounder of the exposure-outcome relationship. (3) Exclusion restriction – G is independent of the outcome conditional on the exposure and all confounders of the exposure-outcome relationship (i.e., the only path between the instrumental variables and the outcome is via the exposure).*

*Supplementary Figure 3*

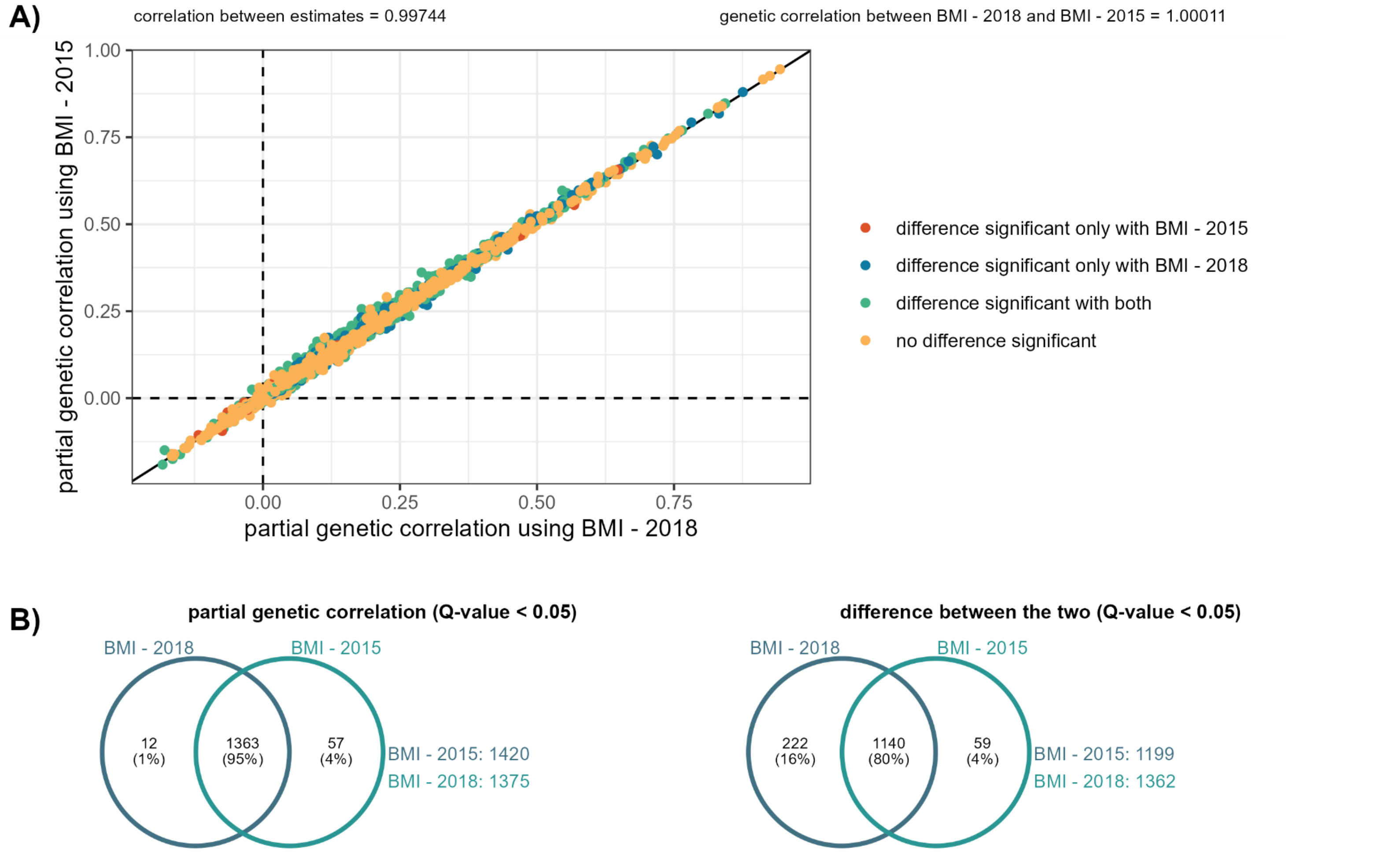


*Comparison of the partial genetic correlation results obtained using BMI data from 2015 and BMI data from 2018. Panel A is a scatter plot of the partial genetic correlation estimates (coloured according to the statistical significance of the difference between the unadjusted and the partial correlation estimates). Panel B presents Venn Diagrams of the statistically significant results for the partial genetic correlation estimates, and the difference between the unadjusted and the partial correlation estimates.*

*Supplementary Figure 4*

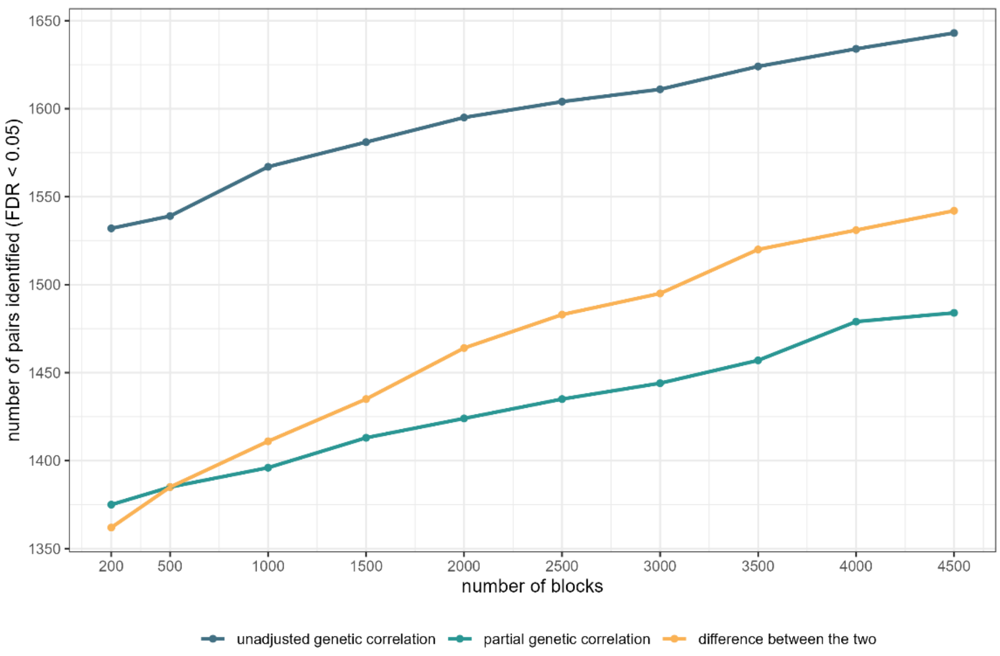


*Effect on the number of blocks used for jackknife on the number of significant signals, for unadjusted genetic correlation (blue), partial genetic correlation (blue-green), and the difference between the two (yellow).*

_Supplementary Figure 5_

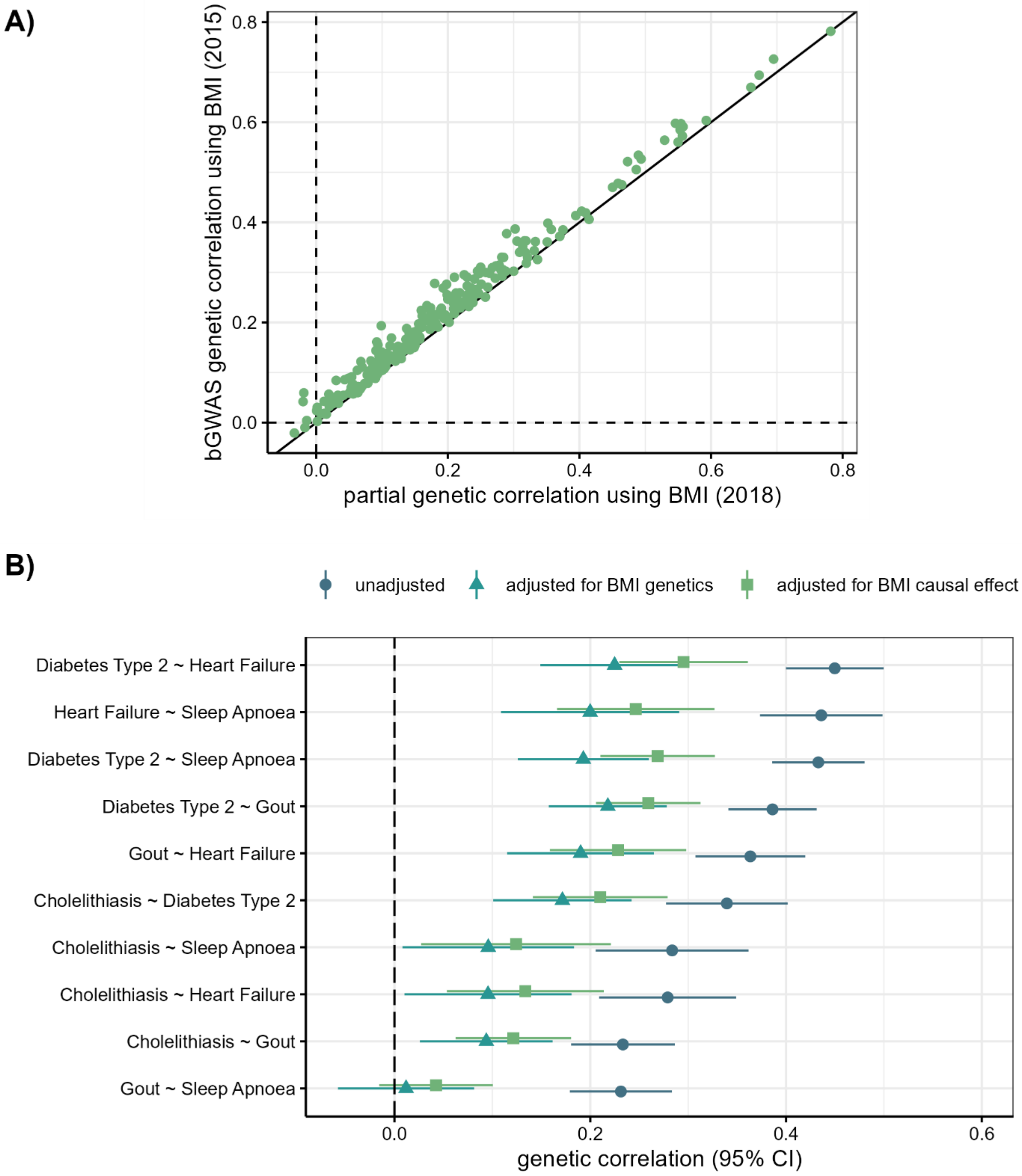


*Panel A: Comparison of partial genetic correlation estimates (using BMI data from 2018, x-axis) and bGWAS genetic correlation estimates (estimated from direct effects obtained using bGWAS and BMI data from 2015, y-axis). Panel B: Unadjusted (blue circle), partial (blue-green triangle) and bGWAS (green square) genetic correlation estimates and 95% confidence intervals for the 10 pairs encompassing the 5 conditions that are the most strongly affected by BMI.*
